# Molecular Detection of *Chlamydia trachomatis* in Infertile Syrian Women: A Comparative Analysis of *ompA* and Cryptic Plasmid PCR Assays

**DOI:** 10.1101/2025.08.05.25333095

**Authors:** Manal Almukdad, Taissir Albouni, Haitham Abbassi

**Affiliations:** Department of Laboratory Medicine, Faculty of Medicine, Damascus University, Damascus, Syria; Department of Obstetrics and Gynecology, Faculty of Medicine, Damascus University, Syria

**Keywords:** *Chlamydia trachomatis*, infertile women, STI, PCR, NAATs, primers, *ompA* gene, cryptic plasmid

## Abstract

**Background:** *Chlamydia trachomatis* is a highly prevalent sexually transmitted infection (STI) strongly associated with female infertility. Polymerase Chain Reaction (PCR) is considered the gold standard for diagnosis, utilizing primers designed to target either the *ompA* gene (encoding the major outer membrane protein) or the cryptic plasmid gene.

**Methods:** This cross-sectional study was conducted on infertile women attending the Infertility Clinic at Damascus University Obstetrics Hospital, Syria. A total of 160 cervical swab samples were analyzed from these women. Genomic DNA was extracted using a Qiagen kit, followed by PCR with primers targeting Chlamydia trachomatis major outer membrane protein gene (CTM -*ompA*-targeting) and cryptic plasmid (CTP -cryptic plasmid-targeting).

**Findings:** Positivity rates were 51.3% for CTM and 31.9% for CTP. A statistically significant association was observed between *C. trachomatis* infection and monthly income (p= 0.039), as well as between it and education level (p= 0.001). Additionally, a significant association was found between *C. trachomatis* positivity and education level (p= 0.017).

**Interpretation:** These findings underscore the significant burden of *Chlamydia trachomatis* in infertile Syrian women and highlight the diagnostic advantages of *ompA-*based PCR, suggesting socioeconomic factors influence infection prevalence.

## Introduction

*Chlamydia trachomatis* is an obligate intracellular bacterium, categorized within the family Chlamydiaceae, order Chlamydiales, phylum Chlamydiae, and genus *Chlamydia*. Beyond C. trachomatis, this genus encompasses ten other species: *C. pneumoniae, C. pecorum, C. felis, C. psittaci, C. abortus, C. caviae, C. gallinacea, C. avium, C. suis, and C. muridarum*. Each of these species exhibits distinct natural hosts (animal or human) and tissue tropisms (1). Globally, C. trachomatis is recognized as the most frequently diagnosed bacterial sexually transmitted infection (STI) (2). Among the chlamydial species, C. trachomatis is particularly significant for human infections, as it exclusively infects humans and is linked to ocular and anogenital infections, with the latter being sexually transmitted (1).

C. trachomatis is an obligate, intracellular, Gram-negative bacterium. Its serovars, numbering 19, are identified based on specific epitopes encoded by the *ompA* gene, which produces the major outer membrane protein (MOMP). Serovars A to C are responsible for trachoma, while D to K are associated with urogenital, ocular, and rectal infections. Serovars L1 to L3 are implicated in lymphogranuloma venereum (LGV) (3).

Intracytoplasmic inclusion bodies within *Chlamydia trachomatis* contain glycogen, and these bacteria are susceptible to inhibition by sulfonamides. They are known to cause eye and genital infections in adults, and conjunctivitis and pneumonia in infants (4). Clinically, C. trachomatis typically manifests as cervicitis in women and epididymitis and infertility in men. It’s important to note that infections can often be asymptomatic in both sexes. Azithromycin is a key consideration for treatment (4). In 2020, the World Health Organization reported an estimated 128.5 million new *Chlamydia trachomatis* (CT) infections globally among adults aged 15-49 (5).

C. trachomatis propagates via a distinctive two-day developmental cycle occurring within human or other eukaryotic host cells (6). The chlamydial infectious cycle generally spans 48 to 96 hours. It begins with elementary bodies (EBs), which are approximately 250 nm in diameter and appear as electron-dense cocci. EBs maintain a compact morphology, facilitated by chromatin condensation mediated by histone-like proteins HctA and HctB (7). These infectious, yet transcriptionally inactive, EBs attach to host cells and are subsequently internalized into a membrane-bound vacuole, known as an inclusion. Upon internalization, EBs differentiate into noninfectious, transcriptionally active reticulate bodies (RBs). RBs then divide and synthesize bacterial effectors. As the inclusion develops, chlamydial effectors are secreted into the host cell’s microenvironment and onto the surface of the inclusion membrane. Here, these bacterial effectors manipulate the host cell to safeguard the inclusion, a critical step in regulating C. trachomatis pathogenicity and survival (8). As the cycle progresses, RBs asynchronously differentiate back into EBs (7). Conventional cell culture, once considered the gold standard for C. trachomatis detection, is no longer practical for rapid diagnostic demands due to its prolonged culture cycles and the need for highly skilled technicians (5). Presently, molecular typing methods are employed for genotyping, which involves analyzing the *ompA* gene that codes for the major outer membrane protein (MOMP); this eliminates the requirement for cell line cultivation. Various genovars have been named using similar letter-based nomenclatures as for serovars. Some of the typing methods utilized include *ompA* sequencing, multilocus sequence typing (MLST), multilocus variable number tandem repeat analysis (MLVA), and whole -genome sequencing (WGS) (9). Given that Chlamydia is an obligate intracellular bacterium, specimens collected for molecular detection must contain host cells harboring the pathogen, especially when direct methods are utilized for testing (9).

Genital swabs are the most frequently collected specimens for detecting chlamydial infections, with Dacron, rayon, cotton, and calcium-alginate tipped swabs generally preferred. In women, cervical swabs, or a combination of cervical and urethral swabs, are considered effective for Chlamydia isolation (9).

The primary challenge in diagnosing human genital Chlamydia infection stems from its often asymptomatic nature or the absence of specific clinical signs. If left undiagnosed, this infection can progress to severe reproductive complications in affected individuals, including pelvic inflammatory disease (PID) and tubal infertility (10). Moreover, untreated infections can lead to serious and irreversible long-term consequences in women, such as tubal infertility, chronic pelvic pain, and ectopic pregnancies, all contributing to high medical costs and significant psychological distress for patients (3). Traditional diagnostic approaches for C. trachomatis (CT) infections present several drawbacks, including low sensitivity, the necessity for invasive specimen collection, extended turnaround times for performance and reporting, and associated high costs. Furthermore, these tests are prone to false-negative results, which can exacerbate infection spread and increase the incidence of complications. These limitations underscore the urgent need for developing diagnostic tests with enhanced sensitivity and specificity, whether for use as standalone reference tools or in conjunction with established traditional methods. The advent of molecular methods has significantly advanced CT diagnosis, facilitating improved case identification and management (9).

C. trachomatis possesses a remarkably conserved genome, approximately 1.05 megabases (MB) in size, exhibiting high identity in both sequence and gene order. C. trachomatis strains are conventionally typed based on the Major Outer Membrane Protein (MOMP) sequence, encoded by the *ompA* gene. This protein contains four variable domains, and differences in these sequences allow for the serovar grouping of various strains (11). The 7.5 kb chlamydial cryptic plasmid remains a widely utilized sequence target for Chlamydia trachomatis nucleic acid amplification tests (NAATs) (12). All patients diagnosed with chlamydial infection must abstain from sexual activity for 7 days post -treatment, and their sexual partners also require testing and treatment as indicated (13).

## Materials and Methods

### Study Participants and Samples

This cross-sectional study was conducted between August 2023 and April 2024. Participants were women attending the Infertility Clinic at Damascus University Obstetrics Hospital, Syria. The study protocol was approved by the Ethics Committee of the Faculty of Medicine at Damascus University. Additional approval for patient recruitment was obtained from the Damascus University Maternity Hospital. Participants were included in the study after providing informed consent, with a clear understanding of the study’s objectives and future implications. Participant anonymity and data confidentiality were strictly maintained by assigning unique identification numbers, which is more precise. Participants were not offered any financial compensation or material benefits for their involvement. They were explicitly informed that their participation would not incur any additional costs for the medical care they receive at the hospital, nor would they be subjected to any additional invasive laboratory tests solely due to their participation.

The sample size was determined using *G*^*\**^*Power* software, resulting in a total of 160 participants. Participants were divided into two groups based on the type of infertility: primary (n=103), and secondary (n=57) infertility. The age of participants ranged from 17 to 45 years, with a mean age (± standard deviation) of 31.14 (± 6.734) years. Participants were categorized into age groups: 17-20 (5.6%), 21-30 (40.6%), 31-40 (45.6%), and 41-45 (8.1%). Inclusion criteria encompassed women diagnosed with infertility who attended the hospital. Exclusion criteria included women in their menstrual period, and women who had received antibiotics within 4 weeks prior to the study. Endocervical swabs were collected from the participants by a specialized physician after obtaining informed consent using Copan eSwab® collection kits. Samples were immediately transported to the Department of Biology Research Lab at the Faculty of Science, Damascus University, and stored at 4°C. For molecular analysis, DNA was extracted from the collected samples using the DNeasy® Blood & Tissue Kit (QIAGEN®) according to the manufacturer’s protocol. The extracted DNA was then stored in the laboratory as frozen aliquots (-20°C). Subsequent amplification was performed using Polymerase Chain Reaction (PCR) with the LifeECO BIOER® system and specific primers from Macrogen® (Table 1).

**Table 1.**
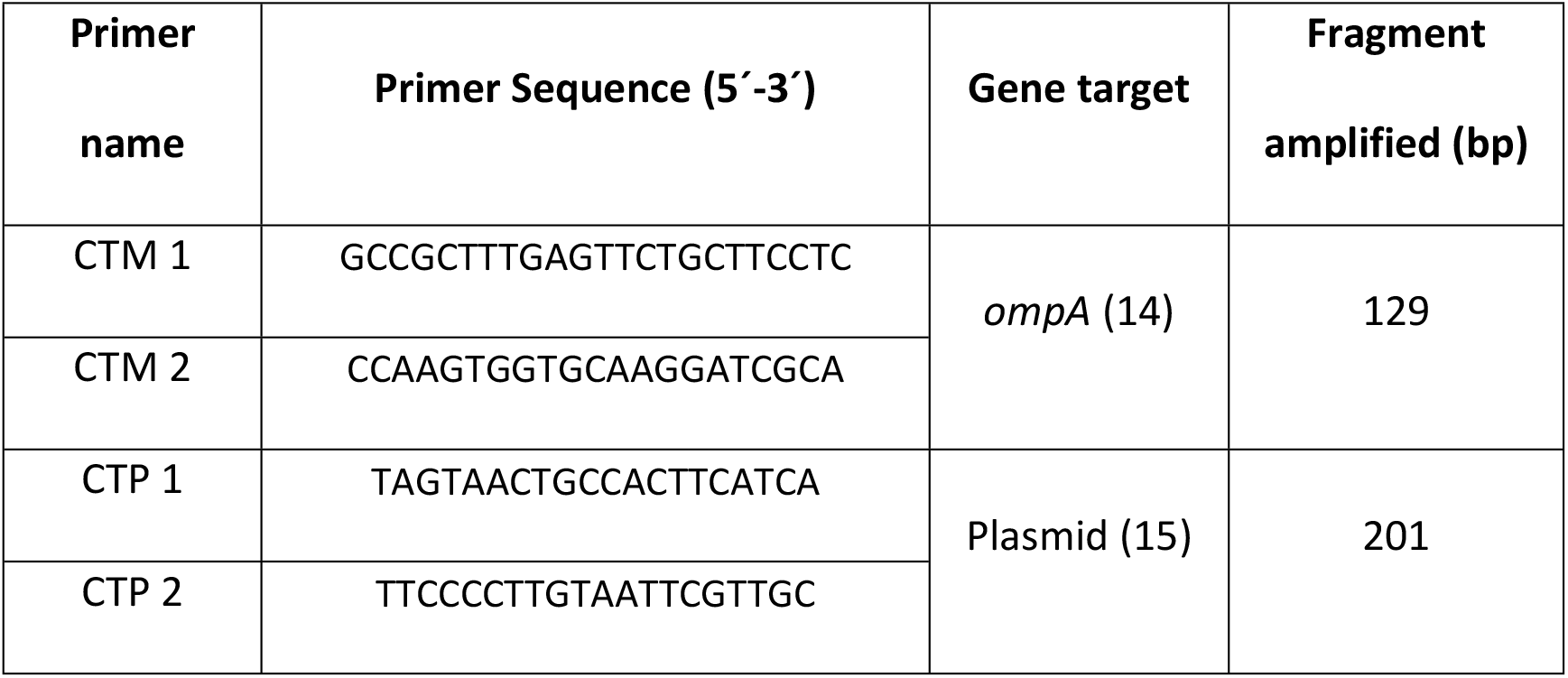
PCR Primer Sequences and Target Information.

The PCR reaction was performed in a final volume of 25 microliters (*μ*L), consisting of 12.5 *μ*L of OnePCR^™^ (GeneDirex®) Ready-To-Use Master Mix containing *Taq* DNA polymerase and deoxyribonucleotide triphosphates (dNTPs). Additionally, 2 *μ*L of genomic DNA from each sample, 2 *μ*L of each primer at a final concentration of 10 picomoles per microliter (pmol/*μ*L), and 6.5 *μ*L of sterile distilled water were added to reach the final reaction volume. The PCR thermocycling conditions were as follows:

### Initial denaturation

94°C for 5 minutes 40 cycles of: Denaturation 94°C for 45 seconds, Annealing: [60°C for 1 minute (for primer CTM), 50°C for 1 minute (for primer CTP)], Extension: 72°C for (1) minute, Final extension: 72°C for 8 minutes.

### Holding 4°C (∞)

PCR amplification products were visualized by electrophoresis on the 2% agarose gel stained with ethidium bromide under Ultraviolet (UV) light. A 100 bp DNA ladder (GeneDirex®) was used to compare the sizes of the amplified products. The resulting amplicon sizes were compared with a known positive control for the PCR reaction obtained from Anatolia Geneworks®, Turkey. A negative control, containing all PCR reagents except the DNA template, was included to ensure optimal reaction conditions and to monitor for contamination.

Statistical analysis was performed using IBM SPSS Statistics version 24 software. The frequency of *Chlamydia trachomatis* positive rates, as detected by both CTM and CTP primers, was calculated, and their respective positive rates were compared. The association of *C. trachomatis* with demographic characteristics was evaluated using Pearson chi-square or Fisher’s exact tests. *p-*value of < 0.05 was established as statistically significant.

## Results

Our study included 160 cervical swab samples from women diagnosed with infertility and who met the inclusion/exclusion criteria. The sample size was calculated using G^*^Power software. Participants’ ages ranged from 17 to 45 years, with the mean age of 31 years old. Descriptive statistics were used to summarize the baseline characteristics of the study cohort. Frequencies and percentages were computed for categorical variables such as age, education level, monthly income, participants’ general health status (hypertension, asthma, diabetes mellitus) (Table 2, Table 3).

**Table 2.** Age Distribution of Study Participants.

| Age Groups (years) | Frequency (n) | Percent (%) |
| --- | --- | --- |
| 17-20 | 9 | 5.6 |
| 21-30 | 65 | 40.6 |
| 31-40 | 73 | 45.6 |
| 41-45 | 13 | 8.1 |
| Total | 160 | 100.0 |

**Table 3.**
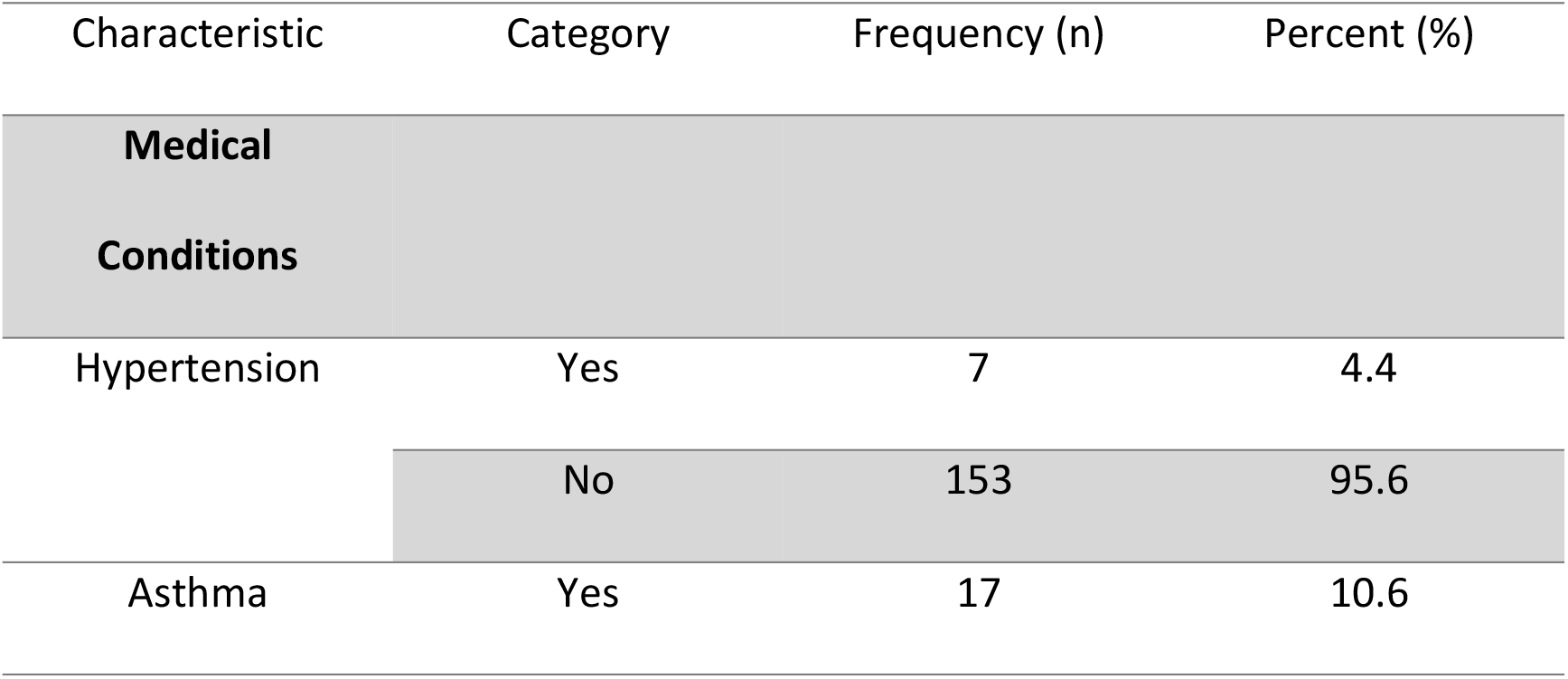
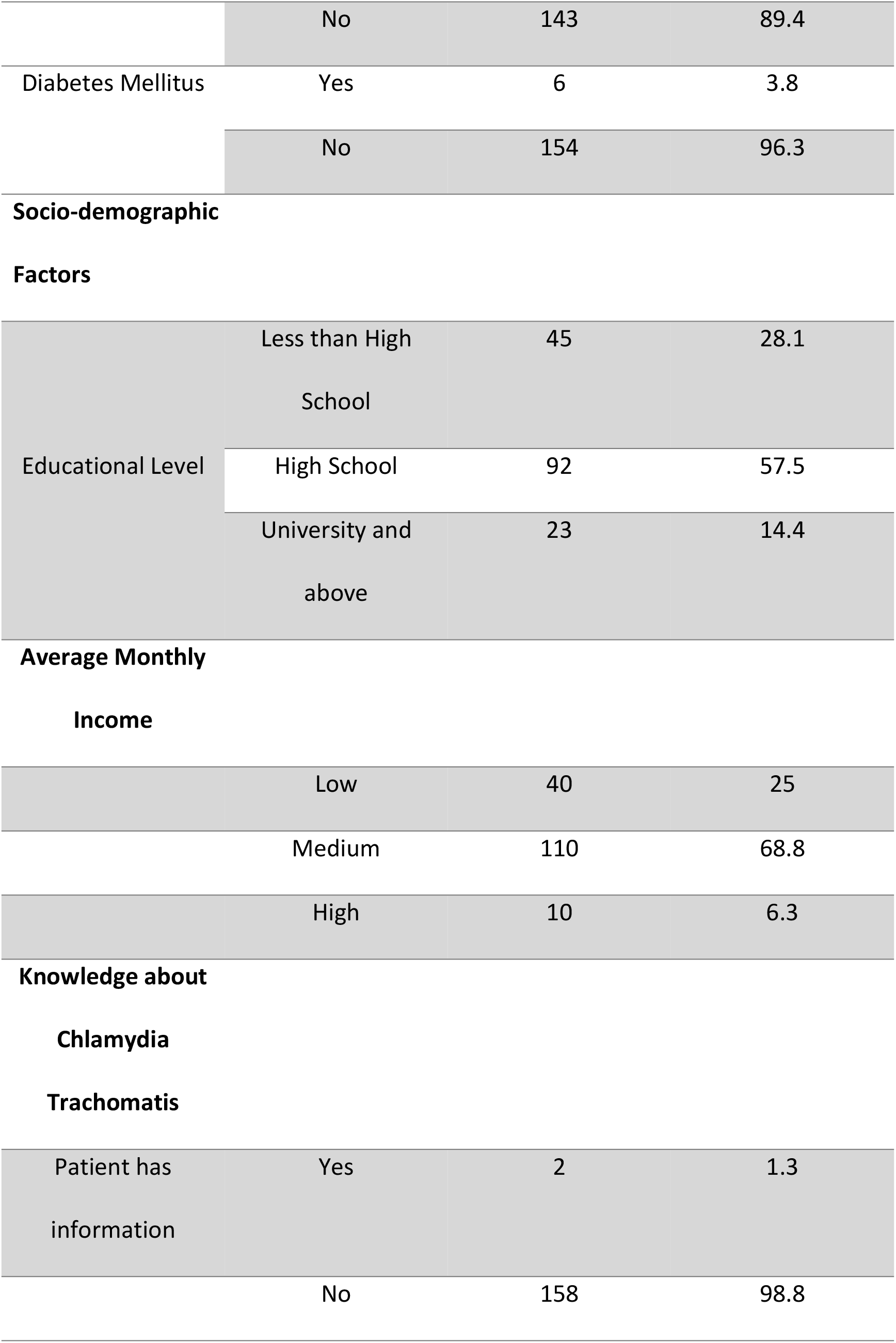
Baseline Characteristics and Health Profile of Study Participants (n= 160)

PCR products were resolved and their lengths compared using a 100 bp DNA ladder. Specific amplicon sizes were used to confirm *Chlamydia Trachomatis* infection. The CTM primer set yielded an amplicon of 129 bp (Figure 1), while the CTP primer set yielded an amplicon of 201 bp (Figure 2), which were the target sizes in this study. The use of these two primer sets aimed to diagnose *Chlamydia trachomatis* strains, including those carrying the cryptic plasmid.

**Figure 1.**
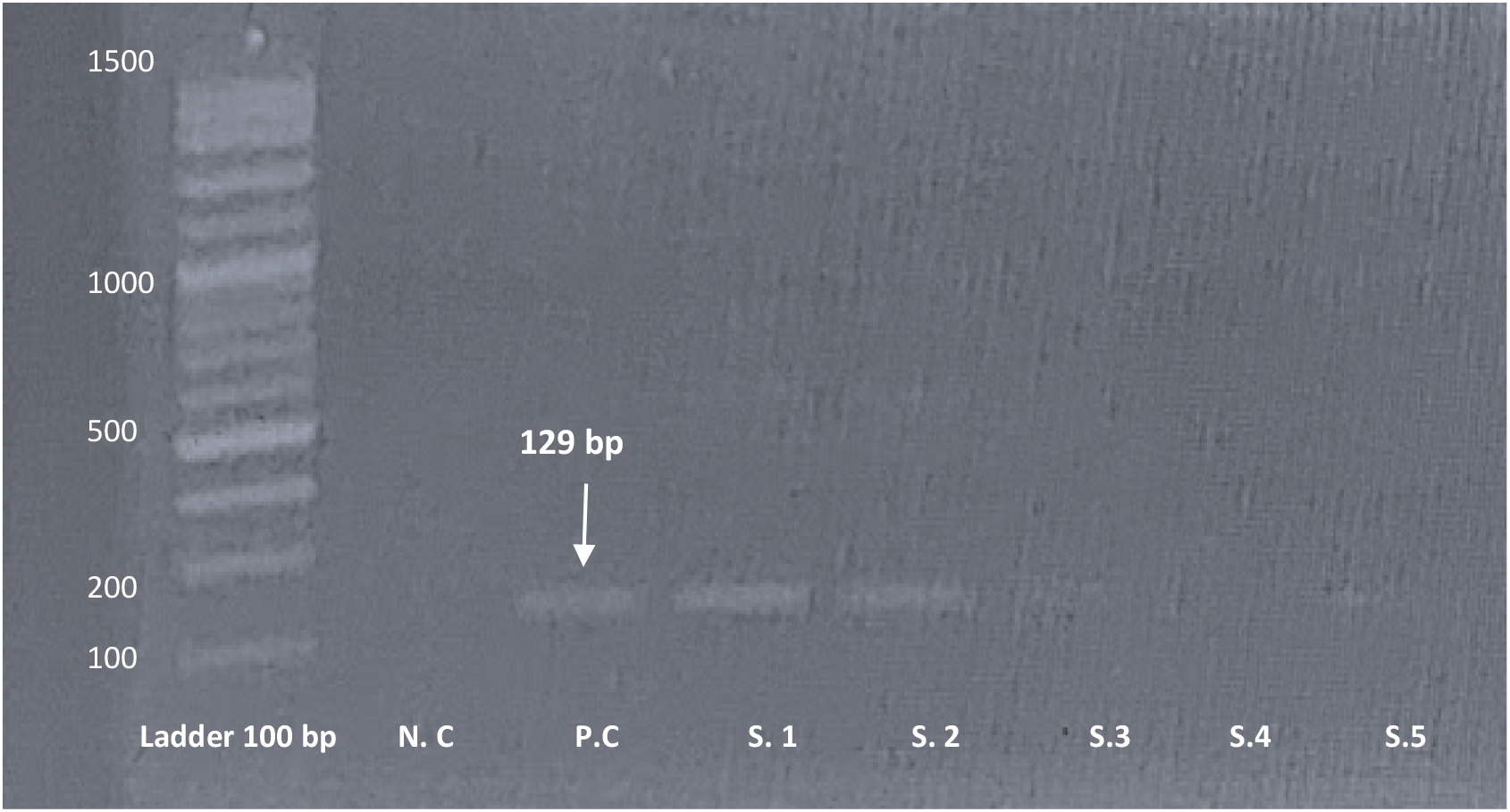
Detection of *Chlamydia trachomatis* DNA in Clinical Samples by PCR using CTM Primers. Agarose Gel Electrophoresis of PCR Products Amplified with CTM Primers. Lane 1 shows a 100 bp DNA ladder. Lane 2 represents the negative control (N.C). Lane 3 shows the positive control (P.C) demonstrating the expected 129 bp amplicon. Lanes 4-8 represent individual patient samples (S.1 – S.5), illustrating positive amplification. The electrophoresis was run on a 2% agarose gel. (**Original work by the authors**).

**Figure 2.**
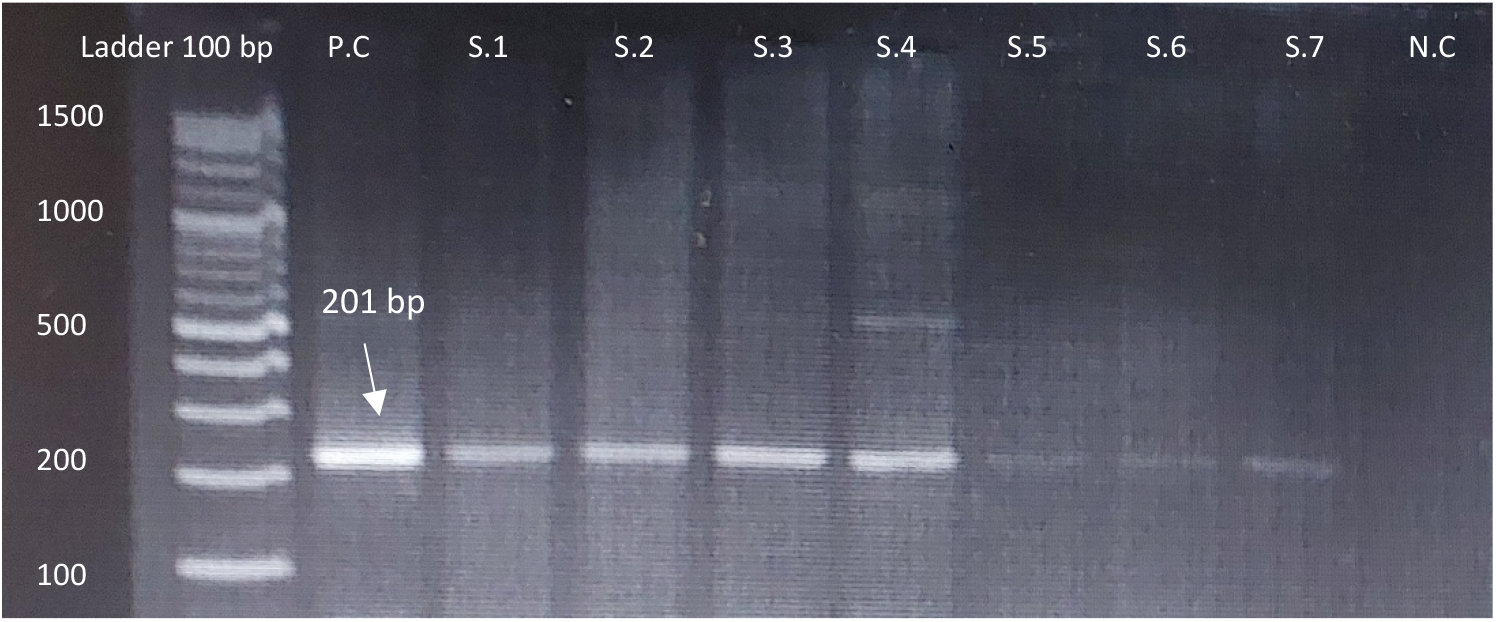
Detection of *Chlamydia trachomatis* DNA in Clinical Samples by PCR using CTP Primers. A 2% agarose gel, stained with ethidium bromide, displaying the amplification products. **Ladder:** Lane ‘Ladder 100 bp’ denotes the 100 base pair DNA ladder. **Positive Control (P.C):** Lane ‘P.C’ exhibits a distinct band at the expected 201 bp size. **Patients’ Samples (S.1-S.7):** Lanes ‘S.1’ through ‘S.7’ represent PCR amplification from individual patients’ samples. **Negative Control (N.C):** Lane ‘N.C’ (Negative Control) confirms the absence of contamination. **Arrow:** The white arrow indicates the 201 bp amplicon corresponding to the CTP target sequence of *Chlamydia trachomatis*. (**Original work by the authors**).

The demographic and clinical characteristics of the study population, along with their association with *Chlamydia trachomatis* positivity (determined by CTM and CTP primer sets), are summarized in Table 4. Chi-square tests were performed to assess these relationships.

**Table 4.**
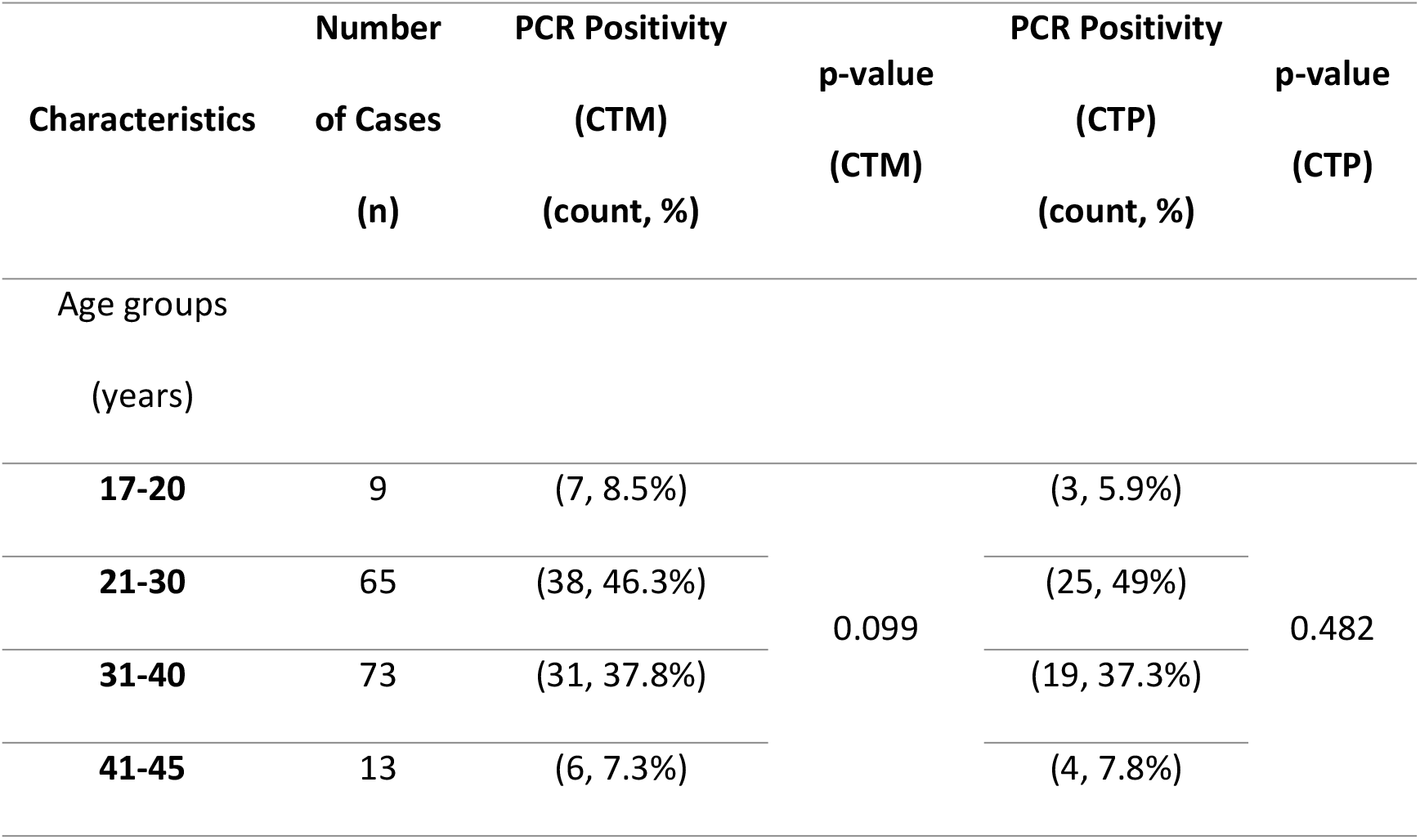
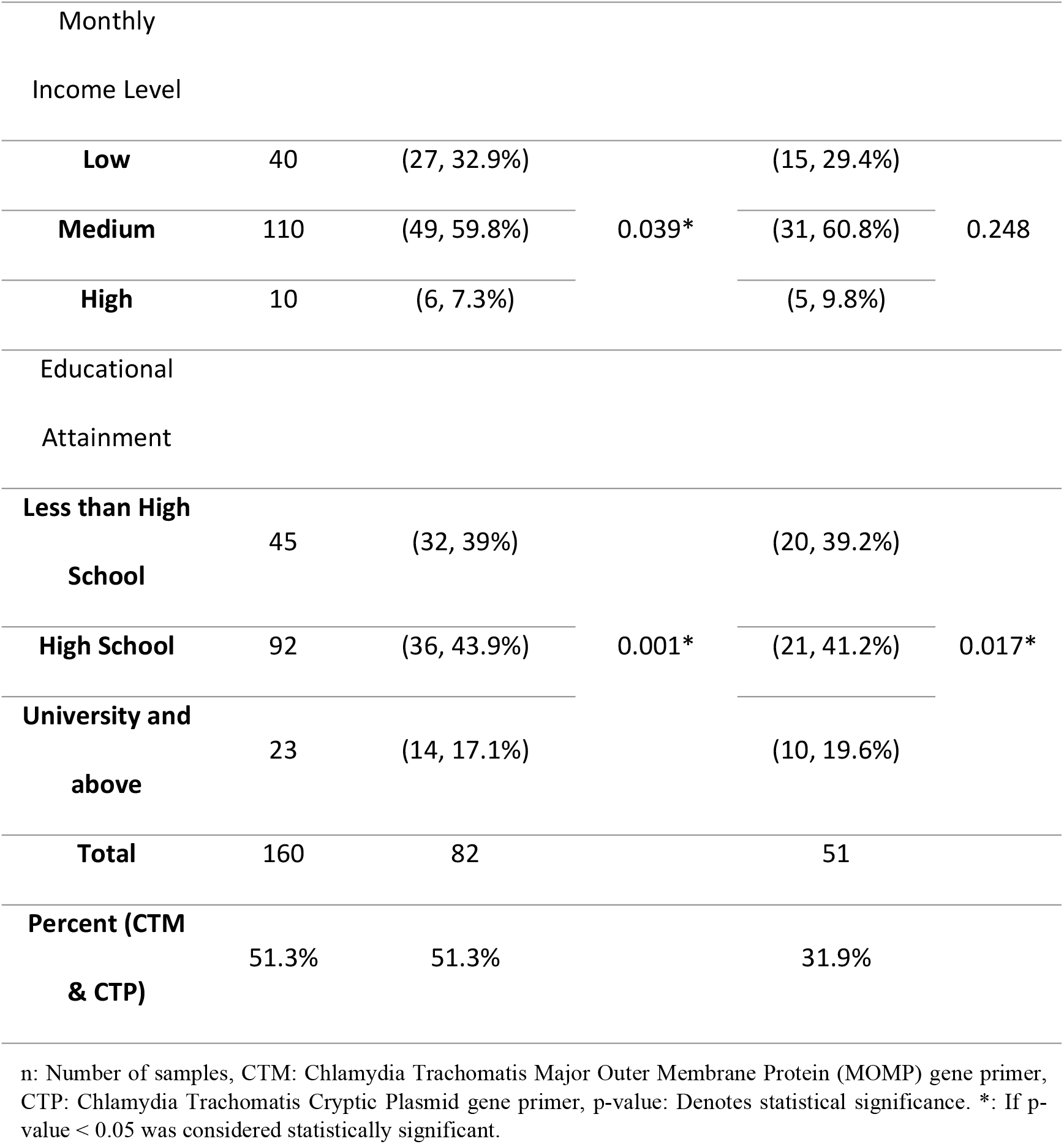
Prevalence of *Chlamydia Trachomatis* Positivity by PCR According to Sociodemographic Characteristics (N = 160)

## Discussion

This study represents a pioneering effort in Syria, as it is the first to employ Polymerase Chain Reaction (PCR) for the detection of *Chlamydia trachomatis*, providing critical insights into its prevalence within a cohort of infertile women. In this study, out of the 160 patients examined, 82 (51.3%) tested positive for *C. trachomatis* infection. A key finding was the statistically significant difference in detection rates between the two primer sets employed, with Primer CTM yielding a substantially higher detection rate of 51.3% (n= 82) compared to 31.9% (n= 51) for Primer CTP (p < 0.001). This disparity suggests that Primer CTM may target a more conserved or abundant genomic region of *C. trachomatis*, thus offering superior sensitivity for molecular detection. The enhanced performance of Primer CTM has significant implications for diagnostic accuracy in routine clinical settings, particularly in resource-limited environments where precise and reliable detection is paramount for effective patient management and public health initiatives. Furthermore, our analysis revealed a compelling association between *C. trachomatis* detection and socioeconomic factors. Specifically, a statistically significant relationship was observed between monthly income level and *C. trachomatis* detection *only* when using Primer CTM. Additionally, a significant association was found between educational attainment and detection rates for both CTM and CTP Primers. These findings underscore the complex interplay of socioeconomic determinants on the epidemiology of *C. trachomatis* infection, highlighting the need for targeted health education and screening programs that address disparities influenced by income and education level. In the context of Syria, a developing and conflict – affected country, the observed relatively high detection rate of *C. trachomatis* among infertile women is consistent with various studies reported from other developing countries. For instance, a 2023 study from Tehran, Iran, involving 291 infertile patients, reported a *C. trachomatis* positivity rate of 10.3% (16). Similarly, a 2023 study from Iraq diagnosed *Chlamydia trachomatis* using real-time PCR in a sample of 125 patients, included 100 infertile women, with a reported detection rate of 12% (17). In Pakistan, a 2024 study on 154 female patients reported a *Chlamydia trachomatis* seroprevalence of 58.4%, detected by enzyme-linked immunosorbent assay (ELISA) for IgG antibodies(18). Among other Arab studies, a 2023 study conducted in Sudan reported a high prevalence of *Chlamydia Trachomatis* (84.6%, 110/130 cases identified via vaginal swabs) and demonstrated a statistically significant association (p<0.05) with age, a finding that contrasts with our observations (19). In Brazil, a study on 498 women showed that the risk factors associated with *Chlamydia trachomatis* were youth (<30 years), no steady sexual partner, co-infection with other sexually transmitted infections (STIs), and low income (20). This aligns with our findings regarding the association between *Chlamydia trachomatis* positivity and low monthly income. In our country, Syria, we observed a high prevalence of *Chlamydia trachomatis* primarily due to a lack of health awareness, inadequate health literacy, and the absence of educational programs concerning the transmission and prevention of sexually transmitted infections. Our study specifically revealed that 98.8% of the participating female patients had no prior knowledge concerning this pathogen (*C. trachomatis)*, its modes of transmission, or associated symptoms. This significant knowledge gap contributes considerably to the wider dissemination of *C. trachomatis* within our community. This is particularly concerning given that approximately 75% of *C. trachomatis* infections are frequently asymptomatic (21).Furthermore, misconceptions about the disease significantly influence individuals’ notions and beliefs regarding its symptoms and prevention. Health officials involved in the management of sexually transmitted diseases (STDs) speculate that adolescents, in particular, may lack awareness of the availability of testing options and the seriousness of *Chlamydia* due to its often asymptomatic nature, especially in the early stages (22). The observed disparity in *Chlamydia trachomatis* diagnostic rates using different primer sets targeting the *ompA* gene (encoding the Major Outer Membrane Protein, MOMP) and the cryptic plasmid gene warrants detailed discussion. In our study, a diagnosis rate of 51.3% was achieved using *ompA*-specific primers (CTM), whereas cryptic plasmid-targeting primers (CTP) yielded a lower rate of 31.9% among infertile women, utilizing cervical swab samples. This significant difference can primarily be attributed to the inherent genetic characteristics of *Chlamydia trachomatis* and the design of the respective diagnostic assays.

The cryptic plasmid, typically present in multiple copies (7-10 copies) per bacterial cell (23), is often targeted by highly sensitive nucleic acid amplification tests (NAATs) due to its high copy number, thereby enhancing detection even with low bacterial loads. Conversely, the *ompA* gene is a single-copy chromosomal target. Therefore, lower sensitivity might be expected when solely relying on *ompA* detection, particularly in samples with limited bacterial DNA. However, our findings, demonstrating that *ompA*- based detection outperformed plasmid-based detection, strongly suggest the prevalence of plasmid-free *Chlamydia trachomatis* variants within our study population. These variants, lacking the cryptic plasmid, would be undetectable by assays exclusively targeting plasmid-borne genes, leading to false-negative results and an underestimation of true infection rates. The existence and clinical significance of plasmid-free *Chlamydia trachomatis* strains are well-documented in the literature, emphasizing the necessity of employing diagnostic strategies that include chromosomal targets to ensure comprehensive detection and avoid misdiagnosis (23). Furthermore, while *ompA* is crucial for serotyping, its high genetic variability across different genotypes could, in some instances, affect primer binding efficiency, potentially influencing detection rates. Our results therefore underscore the importance of considering regional epidemiological variations and the potential circulation of plasmid free strains (24) when selecting and interpreting *Chlamydia trachomatis* diagnostic assays. This is particularly relevant in specific population such as infertile women, where asymptomatic infections are common and can lead to severe reproductive consequences.

Our study has several limitations. Firstly, the relatively small sample size, coupled with the scarcity of previous studies on the prevalence of *Chlamydia trachomatis* in Syria, and the lack of comprehensive Chlamydial typing data (including information on prevalence genotypes within our country), collectively limit the generalizability of our findings. Secondly, being a cross-sectional study, our design precludes definitive conclusions regarding causality. Therefore, future longitudinal studies are warranted to confirm our observations. Additionally, our investigation was confined to Damascus University Obstetrics Hospital, which may impact the broader applicability of these results to other regions or healthcare settings.

## Conclusions and Recommendations

Our study has revealed a concerningly high prevalence of *C. trachomatis* infection within our community, particularly among infertile women. This highlights the crucial role of sexual partners (spouses) as a primary source of infection. Given that Genital tract infections can lead to serious adverse reproductive health consequences for women, these findings underscore the critical importance of early detection and effective antibiotic treatment of *C. trachomatis* infection as a public health priority.

Key recommendations stemming from this research include the necessity for routine screening for *Chlamydia trachomatis*, especially for women experiencing difficulty conceiving. Furthermore, community awareness campaigns emphasizing the importance of preventing sexually transmitted infections and adhering to safe sexual practices are crucial. Implementing comprehensive national screening programs for women to detect preventable diseases and mitigate their associated complications and burden is also vital.

From a microbiological perspective, we recommend conducting more extensive studies and research to confirm the causal relationship between *C. trachomatis* and infertility in our community. Specifically, further molecular investigations and genotyping are needed to identify the prevalent strains in our country and their respective prevalence rates. It is noteworthy that the detection rate for *C. trachomatis* was significantly higher when utilized a primer targeting the *ompA* gene, which encodes the major outer membrane protein. This suggests that *ompA* -targeting primers may be considered more reliable than primers targeting the cryptic plasmid gene for diagnostic purposes.

## Data Availability

Data cannot be shared publicly because of patient confidentiality. Data are available from the Damascus University Institutional Review Board / Ethics Committee (contact via) for researchers who meet the criteria for access to confidential data.

## Ethics statement

The study protocol was approved by the Ethics Committee of Faculty of Medicine at Damascus University, Damascus. (Approval No.: MD-300625-476). Informed consent was obtained from all participants prior to their involvement in the study.

## Abbreviations

CT: *Chlamydia Trachomatis;*
EBs: Elementary Bodies
HctA and HctB: histone-like proteins
MB: Megabases
MLST: Multilocus Sequence Typing
MLVA: Multilocus Variable number tandem repeat Analysis
MOMP: Major Outer Membrane Protein
NAATs: Nucleic Acid Amplification Tests
ompA gene: outer membrane protein A gene
PCR: Polymerase Chain Reaction
RBs: Reticulated Bodies
STI: Sexually Transmitted Infection
WGS: Whole-Genome Sequencing
WHO: World Health Organization.

